# Depression: an adaptive disease?

**DOI:** 10.1101/2023.03.20.23287500

**Authors:** Shiyu Yang, Chenqing Zheng, Canwei Xia, Langyu Gu

## Abstract

Whether depression can be adaptive has long been discussed, but systematically investigation of the adaptive evolution of its genetic basis at the genomic level is sparse. Here, we conducted genome-wide analyses on 320 depression genes at two levels, i.e., across the primate phylogeny (large timescale selection), and in modern human populations (recent selection). We identified seven genes under positive selection in the human lineage, and 46 genes under positive selection in modern human populations. Most positively selected variants in modern human populations were at UTR regions and non-coding exons, indicating the importance of gene expression regulation in the adaptive evolution of depression gene networks. Positively selected genes are not only related to immune response, but also function in reproduction and dietary adaptation, extending the pathogen host defense hypothesis. Notably, the proportion of positively selected depression genes was significantly larger than the proportion of positively selected genes at the genomic level in certain modern human populations. We also identified two positively selected variants that happened to be depression-associated variants. We thus propose that recent selection plays important roles on the adaptive evolution of depression genes. Depression can be adaptive, or is a by-product of evolution, which is still an open question.

## Introduction

Depression ranks as the leading cause of disability and seriously affects the normal life of many people [1]. According to its typical clinical features, depression has multiple negative effects on human beings. The International Classification of Diseases (ICD-10-coded) defined depression as a mental disease in which *the patient suffers from lowering of mood, reduction of energy, and decrease in activity. (Capacity for enjoyment, interest, and concentration is reduced, and marked tiredness after even minimum effort is common. Sleep is usually disturbed and appetite diminished. Self-esteem and self-confidence are almost always reduced and, even in the mild form, some ideas of guilt or worthlessness are often present. The lowered mood varies little from day to day, is unresponsive to circumstances and may be accompanied by so-called “somatic” symptoms)* (https://icd.who.int/browse10/2016/en#/F32). Recent studies have also reported that most suicide subjects have been diagnosed with depression [2]. In general, depression has considerable negative consequences, including notable costs to survival and fitness.

Although depression considerably impairs fitness, a few hypotheses propose that depression can have survival benefits. Some hypotheses focus on the phenotype of depression itself, emphasizing that emotions can affect physiological reactions and behavior, playing adaptive roles in social benefits, such as to solve social problems by rumination [3], to reduce costly conflict, and to avoid infection [4]. But relative evidence is sparse. Other hypotheses emphasize the associations between depression and immune functions, such as the pathogen□host defense hypothesis [5], which proposes that depression can help energy conservation and reallocation to enhance immune response. Indeed, this has been supported by a few studies [6,7]. But there are also findings that cannot be explained. For example, depression exists in the population with low pathogenic exposures [8]. In fact, in addition to immune functions, there may also other fitness related functions that are involved in the depression gene networks due to genetic pleiotropic effects. Without a systematically investigation of depression genes at the genomic level, we will not get a comprehensive picture of gene networks involved in depression. Besides, to what extent that depression genes are under positive selection is also largely unknown.

Natural selection is the major driver of the adaptive evolution of human beings, and selection pressures can occur at different stages during evolution. On the one hand, selection at the species level must be considered on a large timescale, i.e., millions of years and usually worked on the decisive phenotypes for speciation. For example, brain development and walking upright have been reported under positive selection in the human lineage during primate evolution [9,10]. On the other hand, migration out of Africa in the last 100,000 years has further shaped different modern human populations under different selection pressures [11,12], including living environmental changes, diversified food resources, and new pathogens [13]. These recent selection pressures thus work on new genotypes or gene networks that are adaptable. A series of studies have reported that genes involved in local adaptation, such as anti-UV radiation [14], pathogen antagonism [15], and dietary adaptation [16–19] were under positive selection. If these genes were also involved in diseases due to genetic pleiotropic effects, then disease-associated genes can also be under positive selection. Whether this is the case in depression remains unknown.

Growing evidence has shown that depression has a polygenic genetic basis. With the rapid development of next generation sequencing technology, researchers have started to screen depression genes at the whole-genomic level using genome-wide association studies (GWAS). Hundreds of genes were recently identified to be associated with depression by using large cohorts from the UK Biobank, the Million Veteran Program, 23andMe, and FinnGen [20–23]. Next generation sequencing technology also presented well-assembled and annotated genomes of multiple primate species. These findings provide good resources for us to examine the adaptive evolution of depression genes in depth. Therefore, in this study, we evaluated the adaptive evolution of depression genes retrieved from the GWAS studies mentioned above at two different levels, i.e., across the primate phylogeny (large timescale selection, i.e., millions of years) and in modern human populations (recent selection, approximately 100,000 years). Our study will also deepen our understanding of the adaptive evolution of diseases in human beings in a broad sense.

## Methods

### Positive selection detection across the phylogeny

320 depression genes at autosomes identified by GWAS were retrieved from literatures [20–23] (Supplementary File 1). Ensembl IDs of human genes and corresponding orthologous genes of 21 primates across genomes were retrieved from the Ensembl database using BioMart [24]. Only one-to-one orthologous genes with high confidence were analyzed. Coding genes and gff files for each species retrieved from the Ensembl database were used to extract corresponding coding sequences. The longest transcript of each gene was used for further analyses. Codon alignments were conducted using MAFFT implemented in T-coffee [25] with default parameters. An unrooted tree retrieved from the literature [26] was used for PAML analyses (Figure 1).

**Figure 1.**
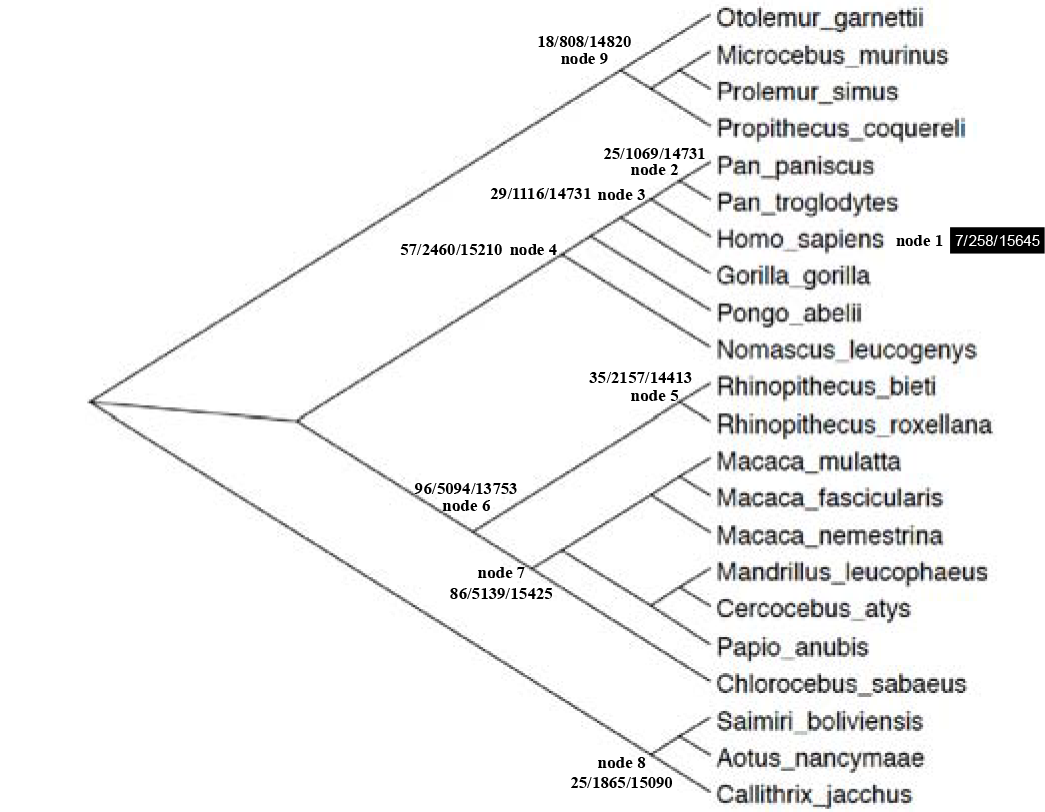
Positive selection detection of depression genes across the primate phylogeny under the branch-site model implemented in PAML. Nine lineages were tested. Numbers on each node from left to the right represent the number of depression genes under positive selection, the number of positively selected genes, and the number of homologous genes retrieved for the PAML test.

The branch-site model in PAML was used to test positive selection across the phylogeny, which consider signals among both lineages and sites [27,28]. Signals of postive selection can be measured by the ratio of nonsynonymous to synonymous substitutions (*ω*). If the *ω* >>1, it means that the nonsynonymous mutations were favored by seletion. We focused on detecting positive selection on nine primate lineages here (Figure 1). To run the branch-site model in PAML, all branches within the target lineage were labeled as foreground branches. Parameters were set as the same as studies that we have published before [29]. Briefly, the M0 model was used to estimate initial branch lengths (fix_blength = 2). Alignment gaps and ambiguity characters were removed (Cleandata=1). The modified ModelA (model = 2, NSsites = 2) and the ModelB (fix_omega = 1, omega = 1) were compared and a likelihood ratio test (LRT) was used to detect significance [27,28]. Multiple test corrections were conducted using Bonferroni correction [30].

### Positive selection detection in modern human populations

Haplotype-based methods (the integrated haplotype score (iHS) and the cross-population extended haplotype homozygosity (xpEHH)) which can limit demographic effects [31–33], as well as pairwise *F*_*ST*_ tests [34] were used to detect positive selection across the genome in different modern human populations, retrieved from the 1000 Genomes Project (ftp.1000genomes.ebi.ac.uk/vol1/ftp/release/20130502/). The iHS test can detect positively selected genes that have not been fixed in the population, and the xpEHH test can detect positively selected genes that near/at fixation [31,35]. To further limit demographic effects, we focused on macro-populations, i.e. the African population, the East Asian population, the European population, and the South Asian population. We also removed recently admixed populations and geographically adjacent populations.

The vcf files of autosomes retrieved from the 1000 Genomes Project were processed with PLINK [36,37] and VCFtools [38]. The GRCh37/hg19 genome was used as the reference. SNPs with indels were not used. Since allele frequency and *F*_*ST*_ are highly correlated, we grouped *F*_*ST*_ values in different allele frequency bins to define the *F*_*ST*_ threshold in each bin, and the top 5% *F*_*ST*_ was used as the threshold. *F*_*ST*_ and allele frequencies were calculated with VCFtools [38]. The selscan software was used to calculate and standardize iHS and xpEHH scores for each SNP across the genome with default parameters [39]. Only biallellic SNPs with minor allele frequency ≥0.05 in test populations were considered. The number of SNPs with |iHS|or |xpEHH|> 2 was counted in a 51-SNP window. Only SNPs with |iHS|or |xpEHH|> 2 in a top 1% 51-SNP window, as well as with high *F*_*ST*_ values were considered under positive selection. We focused on SNPs located in exons in this study. After identifying positively selected genes across the genome in different populations, we compared this gene list with depression genes we have retrieved to identify which depression genes were under positive selection. GO annotations of positively selected genes were conducted with DAVID [40,41].

### Statistical significance tests at the whole genomic level in the human lineage and in modern human populations

Fisher tests were conducted for pairwise comparisons between humans and other primates to determine whether the proportion of positively selected depression genes was significantly different. To determine whether the proportion of positively selected depression genes was significantly larger than the proportion of positively selected genes at the genomic level (at the autosomal level) in each species, we randomly selected 320 genes (the same number as depression genes we tested) across the genome using homologous genes retrieved for PAML analyses each time, and recorded how many of them were under positive selection in the corresponding species. We repeated these steps 100,000 times for each species and plotted the density distribution. Then, we checked whether the actual number of positively selected depression genes was extremely high (located outside the 5% tails of the distribution).

To determine whether the proportion of positively selected depression genes in the target population was significantly larger than the proportion of positively selected genes at the genomic level (at the autosomal level) in that population, we randomly selected 320 genes (the same number as depression genes we tested) across the genome each time and recorded how many of them were under positive selection in that population. We repeated these steps 100,000 times for each population and plotted the density distribution. Then we checked whether the actual number of positively selected depression genes was extremely high (located outside the 5% tails of the distribution). ID and the genome positon of each positively selected variant (the 5’ UTR region, the 3’ UTR region, or the coding region) were retrieved from the Ensembl database.

### Expression quantitative trait loci (eQTLs) for positively selected variants

To see whether positively selected SNPs function as eQTLs, the GTExPortal V8 (https://www.gtexportal.org/home/) was used to acquire corresponding information [42]. The m-value (Posterior probability from MetaSoft) was used to measure the eQTL effects in multiple tissues [43,44]. If m > 0.9, it means that the variant does have an eQTL effect in the tissue.

## Results

### Depression genes under positive selection across the primate phylogeny

The number of depression genes under positive selection, the number of positively selected genes in total, and the number of homologous genes retrieved for PAML analyses in total can be found in Figure 1. Among the 320 depression genes we tested in this study, seven genes were under positive selection in the human lineage (Table 1), including the immune response-related gene STAU1 [45], neurodegeneration-related gene PSEN2 [46], neurological-related gene ANKK1 [47], electron transfer flavoprotein dehydrogenase ETFDH, zinc fingers and homeoboxes 3 ZHX3, neural development-related gene PCDH9 [48], and iron homeostasis-related gene LYRM4 [49].

**Table 1.**
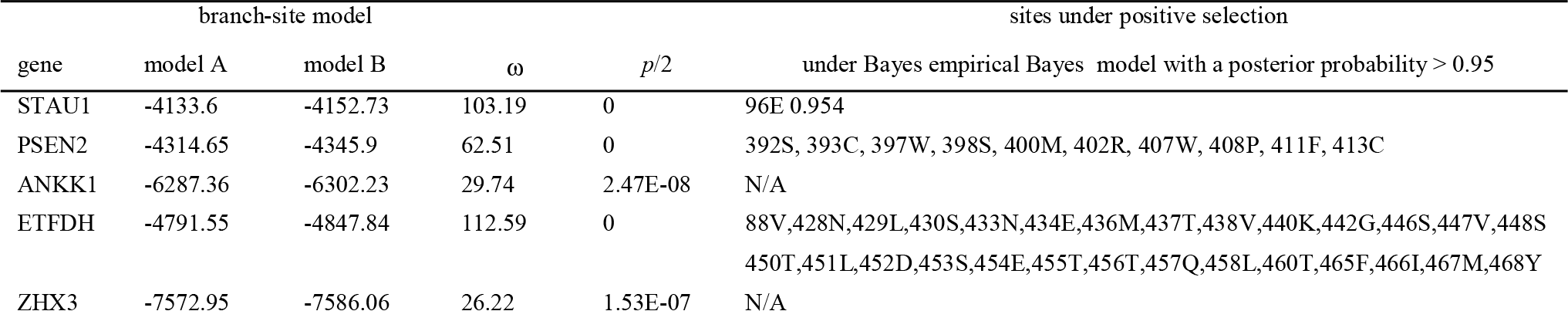
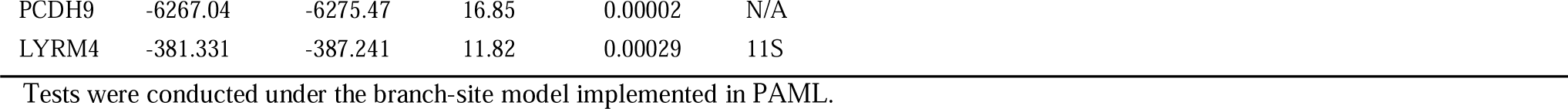
Depression genes under positive selection in the human lineage

Compared to the human lineage, more depression genes were under positive selection in other lineages (Figure 2). For example, 25 depression genes were under positive selection in node 2, which is the sister lineage to human beings, including *Pan troglodyte* and *Pan paniscus*. Twenty-nine depression genes were under positive selection in node 3, the ancestral lineage leading to human beings. Moreover, there were 57 depression genes under positive selection in node 4, 35 depression genes under positive selection in node 5, 96 depression genes under positive selection in node 6, 86 depression genes under positive selection in node 7, 25 depression genes under positive selection in node 8 and 18 depression genes under positive selection in node 9 (Figure 1).

**Figure 2.**
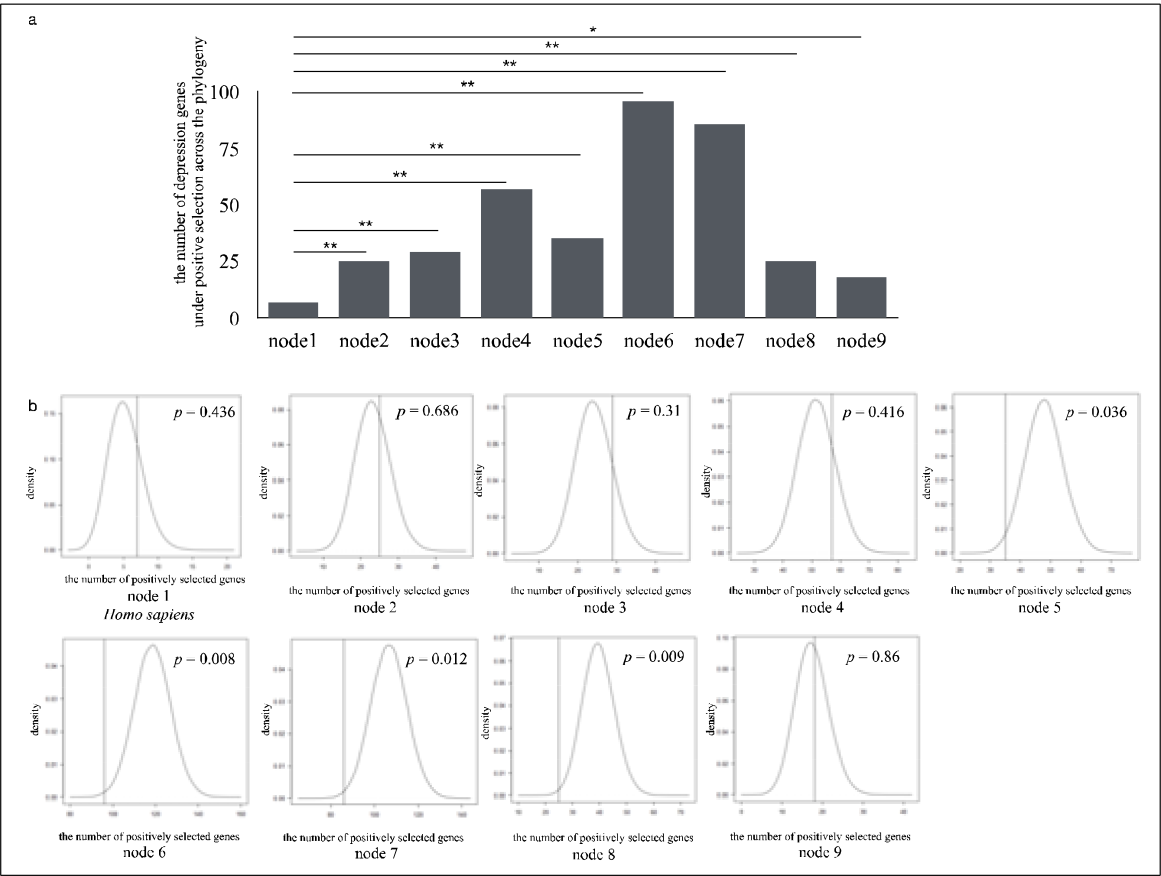
Statistical significance tests of depression genes under positive selection in different primate lineages. (a) Statistical significance tests showed that the numbers of positively selected depression genes in other primate lineages were significantly larger than those in the human lineage. (b) The proportion of positively selected depression genes was not significantly larger than the proportion of positively selected genes at the genomic level. * represents *p* < 0.05; ** represents *p* < 0.01.

We further tested whether the proportion of positively selected depression genes was significantly larger than the proportion of positively selected genes at the whole genomic level. We performed statistical significance tests in all target lineages and did not find any significantly large result. In most cases, there were no significant differences, and sometimes the proportion of positively selected depression genes was even significantly less than the proportion of positively selected genes at the genomic level in certain lineages (node 5, node 6, node 7 and node 8) (Figure 2).

### Depression genes under positive selection in modern human populations

Genome-wide positive selection detection identified 215 genes under positive selection in the African population, 592 genes under positive selection in the East Asian population, 632 genes under positive selection in the European population, and 567 genes under positive selection in the South Asian population. We then identified 46 depression genes under positive selection in total. Nine depression genes were identified under positive selection by the iHS test and the *F*_*ST*_ test. Forty-five depression genes were identified under positive selection by the xpEHH test and the *F*_*ST*_ test (Table 2, Supplementary File 2). Depression genes under positive selection exhibited population-specific patterns. There were eight genes (HLA-DQA1, HLA-DQB1, MGAT4C, ZNF536, SIM1, METTL9, MYRF, PSORS1C1) under positive selection in the African population, five genes (BEND4, CDH13, SOX6, POGZ, PSMB4) under positive selection in the European population, 17 genes (SDK1, CCDC92, DENND1A, USP3, C22orf26, BEND4, ASXL3, TENM2, BSN, ZNF35, MGAT4C, CTNND1, FNIP2, NRD1, INPP4B, C7orf72, FHIT) under positive selection in the East Asian population, and 25 genes (FADS1, FADS2, HLA-DQA1, HLA-DQB1, FAM120A, DENND1A, C22orf26, BEND4, FAM120AOS, ASCC3, LPIN3, POGZ, EMILIN3, CDH9, SGIP1, ZNF660, NICN1, CCDC36, CCDC71, TCAIM, MGAT4C, ZNF445, ZNF197, ZKSCAN7, PHF2) under positive selection in the South Asian population (Table 2, Supplementary File 2). FADS1 and FADS2, which are related to dietary adaptation, have been previously reported to be under positive selection [17–19].

**Table 2.**
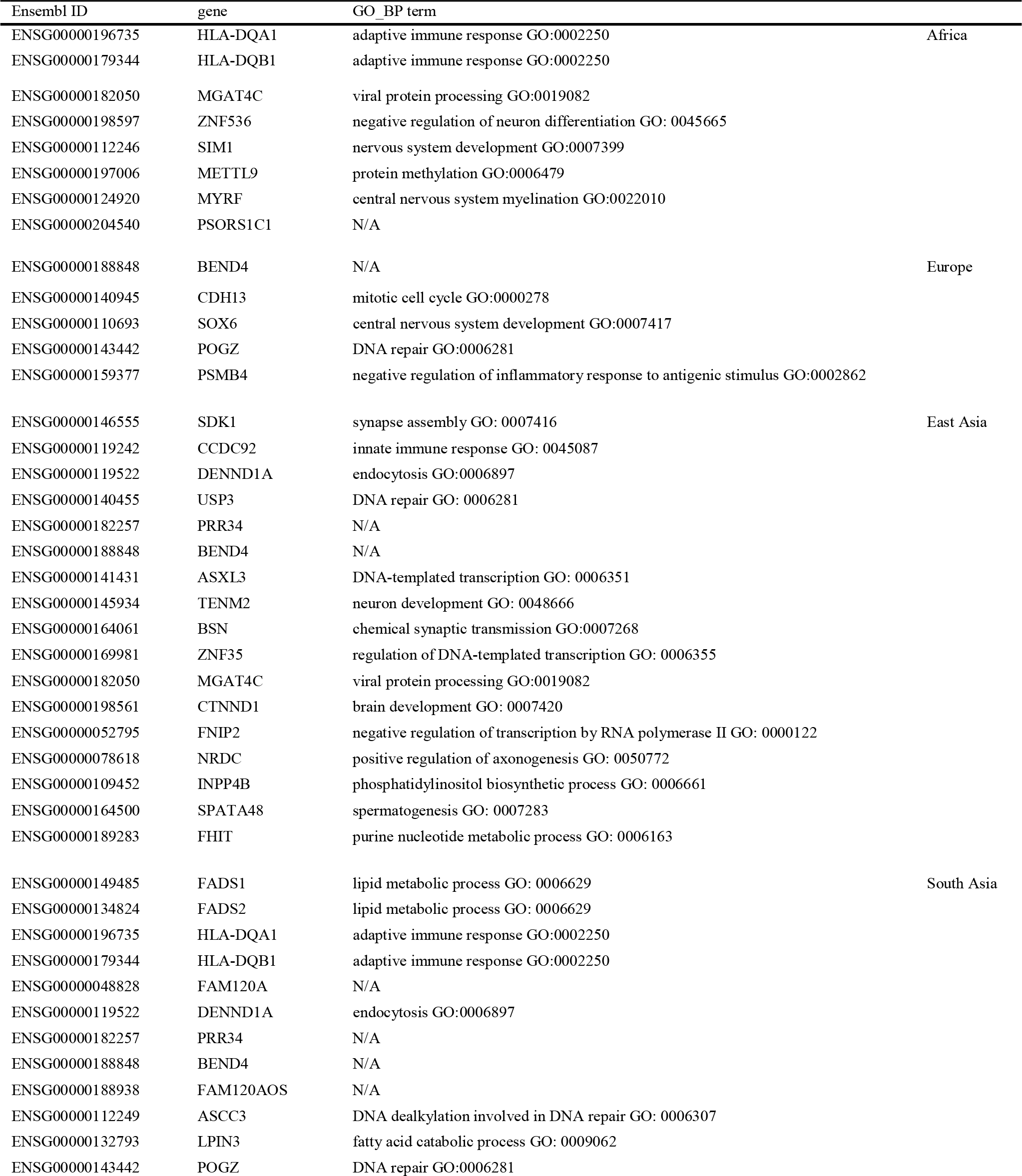
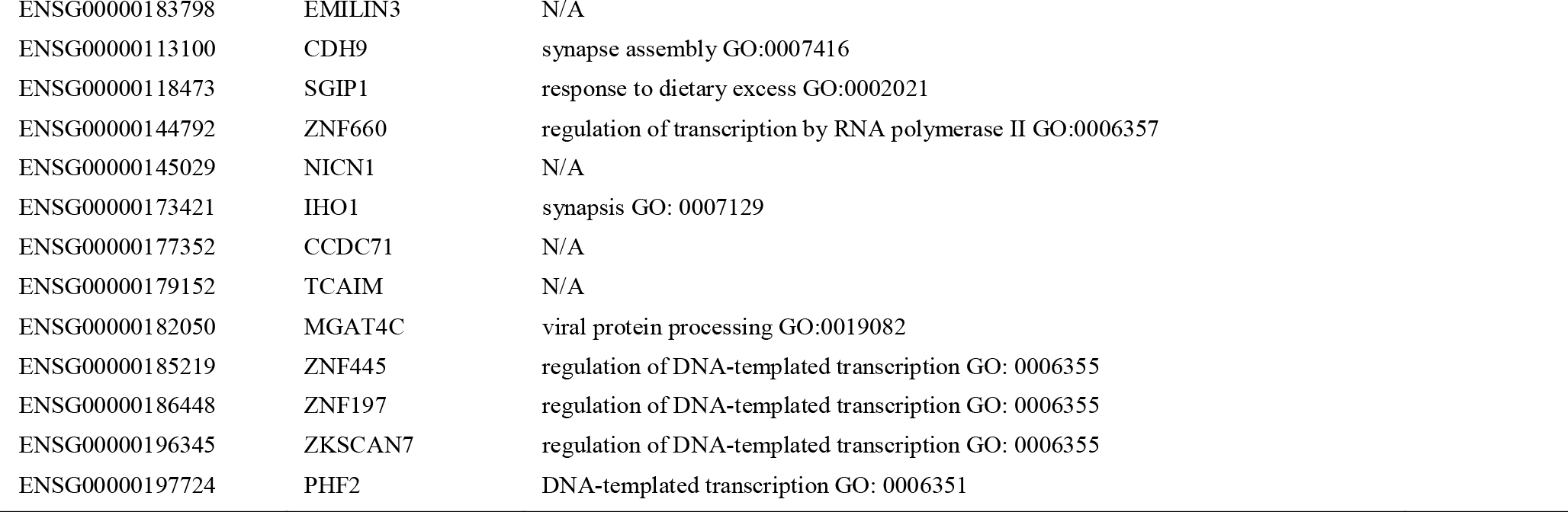
Depression genes under positive selection in modern human populations

| Ensembl ID | gene | GO_BP term |  |
| --- | --- | --- | --- |
| ENSG00000196735 | HLA-DQA1 | adaptive immune response GO:0002250 | Africa |
| ENSG00000179344 | HLA-DQB1 | adaptive immune response GO:0002250 |  |
| ENSG00000182050 | MGAT4C | viral protein processing GO:0019082 |  |
| ENSG00000198597 | ZNF536 | negative regulation of neuron differentiation GO: 0045665 |  |
| ENSG00000112246 | SIM1 | nervous system development GO:0007399 |  |
| ENSG00000197006 | METTL9 | protein methylation GO:0006479 |  |
| ENSG00000124920 | MYRF | central nervous system myelination GO:0022010 |  |
| ENSG00000204540 | PSORS1C1 | N/A |  |
| ENSG00000188848 | BEND4 | N/A | Europe |
| ENSG00000140945 | CDH13 | mitotic cell cycle GO:0000278 |  |
| ENSG00000110693 | SOX6 | central nervous system development GO:0007417 |  |
| ENSG00000143442 | POGZ | DNA repair GO:0006281 |  |
| ENSG00000159377 | PSMB4 | negative regulation of inflammatory response to antigenic stimulus GO:0002862 |  |
| ENSG00000146555 | SDK1 | synapse assembly GO: 0007416 | East Asia |
| ENSG00000119242 | CCDC92 | innate immune response GO: 0045087 |  |
| ENSG00000119522 | DENND1A | endocytosis GO:0006897 |  |
| ENSG00000140455 | USP3 | DNA repair GO: 0006281 |  |
| ENSG00000182257 | PRR34 | N/A |  |
| ENSG00000188848 | BEND4 | N/A |  |
| ENSG00000141431 | ASXL3 | DNA-templated transcription GO: 0006351 |  |
| ENSG00000145934 | TENM2 | neuron development GO: 0048666 |  |
| ENSG00000164061 | BSN | chemical synaptic transmission GO:0007268 |  |
| ENSG00000169981 | ZNF35 | regulation of DNA-templated transcription GO: 0006355 |  |
| ENSG00000182050 | MGAT4C | viral protein processing GO:0019082 |  |
| ENSG00000198561 | CTNND1 | brain development GO: 0007420 |  |
| ENSG00000052795 | FNIP2 | negative regulation of transcription by RNA polymerase II GO: 0000122 |  |
| ENSG00000078618 | NRDC | positive regulation of axonogenesis GO: 0050772 |  |
| ENSG00000109452 | INPP4B | phosphatidylinositol biosynthetic process GO: 0006661 |  |
| ENSG00000164500 | SPATA48 | spermatogenesis GO: 0007283 |  |
| ENSG00000189283 | FHIT | purine nucleotide metabolic process GO: 0006163 |  |
| ENSG00000149485 | FADS1 | lipid metabolic process GO: 0006629 | South Asia |
| ENSG00000134824 | FADS2 | lipid metabolic process GO: 0006629 |  |
| ENSG00000196735 | HLA-DQA1 | adaptive immune response GO:0002250 |  |
| ENSG00000179344 | HLA-DQB1 | adaptive immune response GO:0002250 |  |
| ENSG00000048828 | FAM120A | N/A |  |
| ENSG00000119522 | DENND1A | endocytosis GO:0006897 |  |
| ENSG00000182257 | PRR34 | N/A |  |
| ENSG00000188848 | BEND4 | N/A |  |
| ENSG00000188938 | FAM120AOS | N/A |  |
| ENSG00000112249 | ASCC3 | DNA dealkylation involved in DNA repair GO: 0006307 |  |
| ENSG00000132793 | LPIN3 | fatty acid catabolic process GO: 0009062 |  |
| ENSG00000143442 | POGZ | DNA repair GO:0006281 |  |

|  |  |  |
| --- | --- | --- |
| ENSG00000183798 | EMILIN3 | N/A |
| ENSG00000113100 | CDH9 | synapse assembly GO:0007416 |
| ENSG00000118473 | SGIP1 | response to dietary excess GO:0002021 |
| ENSG00000144792 | ZNF660 | regulation of transcription by RNA polymerase II GO:0006357 |
| ENSG00000145029 | NICN1 | N/A |
| ENSG00000173421 | IHO1 | synapsis GO: 0007129 |
| ENSG00000177352 | CCDC71 | N/A |
| ENSG00000179152 | TCAIM | N/A |
| ENSG00000182050 | MGAT4C | viral protein processing GO:0019082 |
| ENSG00000185219 | ZNF445 | regulation of DNA-templated transcription GO: 0006355 |
| ENSG00000186448 | ZNF197 | regulation of DNA-templated transcription GO: 0006355 |
| ENSG00000196345 | ZKSCAN7 | regulation of DNA-templated transcription GO: 0006355 |
| ENSG00000197724 | PHF2 | DNA-templated transcription GO: 0006351 |

The proportions of depression genes under positive selection in the African population (8 out of 320) (*p* < 0.05), in the East Asian population (17 out of 320) (*p* < 0.05), and in the South Asian population (25 out of 320) (*p* < 0.01) were significantly larger than the proportions of positively selected genes at the genomic level in the corresponding populations (Figure 3).

**Figure 3.**
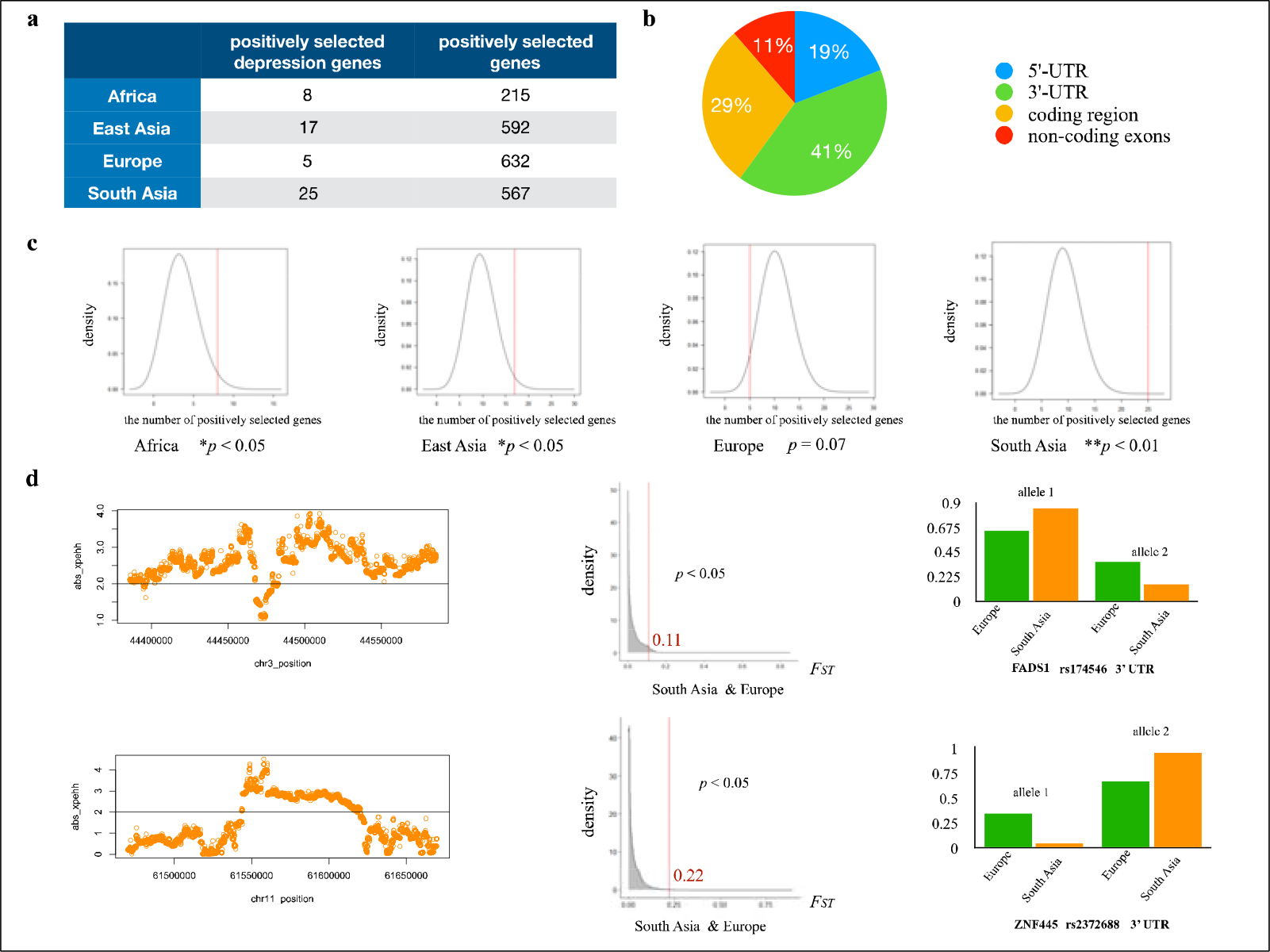
Recent selection plays important roles on the adaptive evolution of depression genes in modern human populations. (a) The number of positively selected depression genes and the number of positively selected genes at the genomic level in each population. (b) Proportions of positively selected variants located in different regions (5’ UTR, 3’ UTR, coding region and non-coding exons). (c) The proportion of positively selected depression genes in the African population, the East Asian population and the South Asian population was significantly larger than the proportion of positively selected genes at the genomic level. (d) left: xpEHH plot of two positively selected variants that happened to be depression-associated variants in the South Asian population compared to the European population. middle: the distribution of *F*_*ST*_ values across the genome between the South Asian population and the European population. The *F*_*ST*_ value of two positively selected variants was located at the top 5% region of the corresponding distribution (*p* < 0.05). right: allele frequencies of two positively selected variants in the European population and in the South Asian population.

The eQTL effects of each positively selected variant at UTR regions were obtained from the GTExPortal V8. Results showed that most positively selected variants have eQTL effects across multiple tissues, and many of them were in non-brain tissues (Figure 4).

**Figure 4.**
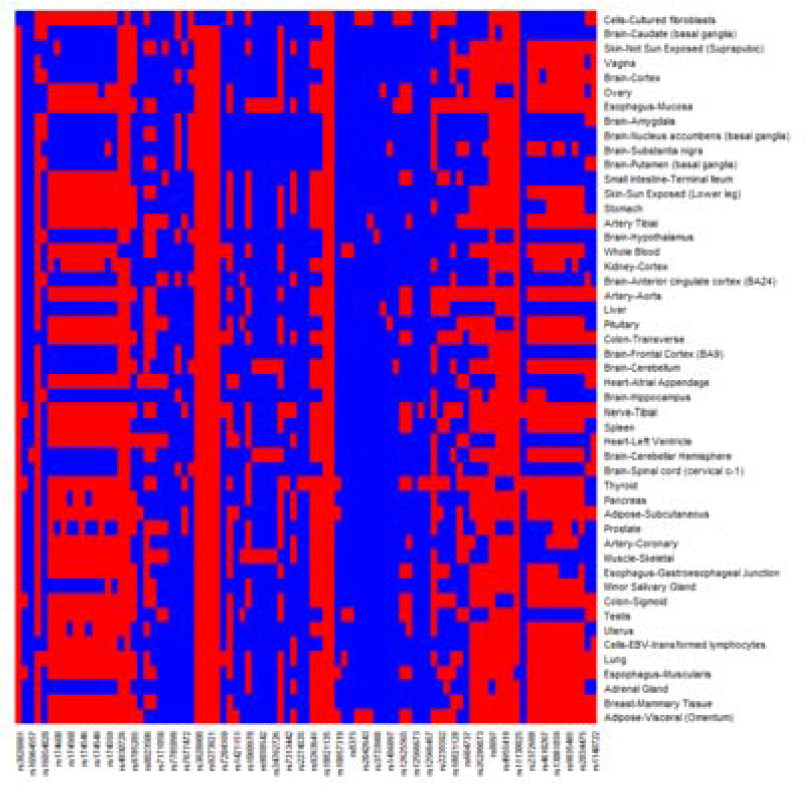
The eQTL effects of each positively selected variant in multiple tissues. The m-value was used to measure the eQTL effects in multiple tissues [43,44]. If m > 0.9, it means that the variant does have an eQTL effect in the tissue. red, m > 0.9; blue, m ≤ 0.9. Only variants with available information in the GTEx database were shown.

## Discussion

We identified 46 depression genes under positive selection in modern human populations, and seven depression genes under positive selection in the human lineage during primate evolution. Many of them are immune-related, consistent with the pathogen□host defense hypothesis that many risk alleles of depression genes are immune-related [5]. Moreover, the SPATA48 gene related to reproduction was identified under positive selection in the East Asian population. There were also positively selected depression genes related to brain or neuron development. Depression as a mental illness, it could be that some of its causative genes are adaptive to cognitive functions in humans, similar to Alzheimer’s disease [50] and schizophrenia [51]. Notably, several dietary adaptation genes were under positive selection in the South Asian population. FADS1and FADS2, the two genes related to the endogenous biosynthesis of long chain polyunsaturated fatty acids that are important to people who rely on plant-based diets, have been reported to be under positive selection in modern human populations to adapt to varied dietary histories [17–19]. In addition, we identified another two genes functioning in the fatty acid catabolic process and dietary excess response under positive selection in the South Asian population, LPIN3 and SGIP1 [52,53]. These results suggest that not only pathogen□host interactions but also other adaptive functions, such as reproduction, cognition development and adaptation to local dietary culture, can play important roles in the adaptive evolution of depression genes. Our study thus extended the pathogen□host defense hypothesis.

This suggests that gene networks involved in depression are adaptive in certain populations under recent selection. The relatively easily acceptable explanation is the genetic pleiotropic effect. Notably, our results showed that most positively selected variants were located in UTR regions and non-coding exons. UTR regions and non-coding exons have been reported to be functioning in gene regulation and were associated with many diseases, including mental diseases [54–57]. The regulation of gene expression is thus important in the adaptive evolution of depression genes under recent selection. Besides, most of them had eQTL effects in multiple tissues, especially in non-brain tissues, confirming their pleiotropic effects involving in multiple biological processes.

It is worth noting that the proportion of positively selected depression genes was so high that it was even significantly larger than the proportions of positively selected genes at the genomic level in certain modern human populations, i.e., in the African population, the East Asian population and the South Asian population. Moreover, we also identified two positively selected variants that happened to be depression-associated variants in two genes, i.e., FADS1 and ZNF445. We thus propose two hypotheses to explain these results, which are not mutually exclusive to each other. One is that depression itself is adaptive in these populations. The classical example that diseases in certain conditions can be adaptive is the sickle cell disease [58]. The red blood cells of carriers of this disease become hard and sticky, but this protects against malaria in Africa. Several hypotheses have already suggest that psychological and behavioural reactions caused by emotion can help people avoid dangerous situations or infections as we mentioned in the Introduction. During human migration in recent history, different populations have to adapt to local conditions. Phenotypes caused by depression can thus be adaptable to local conditions in these populations. The other one is that depression can be a by-product of evolution under recent selection due to pleiotropic effects as we mentioned above.

However, one situation that must be considered before giving a definite answer is that most GWAS studies to identify depression genes mainly used data from individuals of European descent [20–23]. It could be that depression gene-sets are quite different among populations. Recently, one study applied GWAS to identify depression-associated variants in individuals from East Asian descent [59]. They found that only 11% of variants identified from previous studies focusing on individuals of European descent were confirmed in the analysis of individuals of East Asian descent. Although they did not conduct further depression-associated gene-set analyses, which may be due to the limited power of their sample size, their study suggests that part of the genetic basis of depression can be different among human populations. Further GWAS studies to reveal depression genes in non-European populations are thus needed.

One possibility is that the depression gene-sets are quite different in non-European populations compared to the European population, but the proportion of positively selected depression genes was still significantly larger than the proportion of positively selected genes at the genomic level. This will still support our hypotheses. The other possibility is that the depression gene-sets are quite different in non-European populations compared to the European population, and the proportion of positively selected depression genes was not significantly larger than the proportion of positively selected genes at the genomic level. But the phenomenon of population-specific depression genetic basis is still driven by recent selection and local adaptation. And it will not change our findings that genes associated with depression in one population can be adaptive in other populations. We thus propose that recent selection plays important roles on the adaptive evolution of depression genes. Whether depression itself can be adaptive, or is a by-product of evolution under recent evolution, is still an open question.

## Data Availability

All data produced in the present work are contained in the manuscript

## Authors Contributions

SY and LG designed the study. SY and LG conducted positive selection detection in modern human populations. SY, CZ and LG conducted positive selection detection across the phylogeny. SY, CX and LG conducted statistical significance tests. SY and LG wrote the manuscript and all authors commented on it.

